# Healthy Diet is Central in the Network of Health Behaviors among Young Chinese Adults

**DOI:** 10.64898/2026.02.05.26345623

**Authors:** Sicong Liu, Dongshi Wang, Zijian Zhao, Fengwei Hao, Likun Ge, Gaoxia Wei

## Abstract

**Background:** Health behaviors established during young adulthood significantly shape the long-term risk of non-communicable diseases and mental health disorders. Although behaviors such as diet, physical activity, sleep, and substance use are often targeted individually, growing evidence suggests these behaviors function as an interconnected system. However, the organization of lifestyle behaviors at the system level, and which behaviors exert the greatest structural influence, remains poorly understood, particularly in non-Western populations.

**Purpose:** This study aimed to model the interdependence of lifestyle behaviors among university students in China and identify key behaviors with the greatest structural influence within a lifestyle network.

**Methods:** We analyzed cross-sectional survey data from 5,652 university students in China, assessing seven lifestyle behaviors (diet, physical activity, sleep quality, social engagement, green and blue space exposure, alcohol use, and tobacco use) as well as symptoms of anxiety and depression. A pairwise Markov random field model was used to construct a lifestyle network and identify behavioral clusters and influential behaviors. Network stability and subgroup invariance were evaluated using bootstrap and permutation procedures.

**Results:** Three stable behavioral clusters were identified: (1) a positive lifestyle cluster (diet, physical activity, social engagement, and environmental exposure), (2) a distress–sleep cluster (sleep problems, anxiety, and depression), and (3) a substance-use cluster (alcohol and tobacco use). Dietary behavior consistently emerged as the most central behavior in the network, with extensive connections to both behavioral and psychological domains. Physical activity played a more peripheral role. Strong coupling between sleep problems and emotional distress was observed, consistent with systems theories of mental health.

**Conclusions:** These findings support a systems-based framework for understanding health behaviors in young adulthood. Identifying structurally influential behaviors, particularly dietary behavior, can provide leverage points for targeted health interventions. The study highlights implications for public health policy and intervention design, particularly in non-Western university populations.

## Introduction

Noncommunicable diseases (NCDs) represent a dominant and growing public health burden in China, accounting for nearly 70% of total health expenditures and contributing substantially to catastrophic health costs at the household level. These trends underscore the urgency of identifying leverage points for prevention early in the life course, particularly during young adulthood when long-term health trajectories begin to consolidate ^1,2^. The university years constitute a critical developmental window marked by increasing autonomy, identity formation, and exposure to new social and environmental contexts. In China alone, over 47 million individuals are enrolled in higher education in 2023 ^3^, and the college enrollment rate among young adults reached 60.8% in 2025 ^4^, making college students a population of both demographic scale and strategic importance for health promotion.

A defining feature of health behavior is interdependence. Dietary habits, physical activity, sleep quality, substance use, and social engagement rarely develop in isolation. Instead, they co-occur in patterned constellations, shaped by shared contextual constraints (e.g., academic schedules, campus food environments), psychosocial demands, and self-regulatory capacities ^5^. For instance, both diet and physical activity have been linked with sleep patterns in adolescents and young adults ^6,7^, and campus-relevant greenspace/blue-space exposures are associated with better mental health and higher activity levels ^8,9^. Consistently, epidemiological evidence has that clusters of behaviors—rather than any single behavior—more strongly predict morbidity, mental health outcomes, and mortality risk ^10^. This has motivated a shift away from single-behavior models toward systems-oriented approaches that conceptualize lifestyle as a dynamic network of mutually reinforcing practices ^11^.

Several theoretical frameworks converge on this systems-oriented view of health behavior change. Ecological models of health behavior conceptualize individual actions as embedded within nested and interacting layers of influence, including physical environments, social networks, institutional structures, and broader cultural norms ^12^. From this perspective, daily behaviors are not isolated choices but components of a dynamic behavioral ecology, such that perturbations in one domain, such as changes in food availability, social eating norms, or time-use constraints, can cascade across other domains of everyday life. Complementing this view, self-regulation and habit-formation theories emphasize that behaviors differ systematically in their temporal frequency, contextual stability, and cognitive demands ^13^. Behaviors such as eating are enacted multiple times per day, are tightly linked to recurring environmental cues, and often become automatized through habit learning, whereas behaviors such as structured exercise are comparatively episodic, resource-intensive, and more dependent on sustained motivation and executive control ^14,15^. These asymmetries suggest that certain behaviors may function as keystone habits, exerting disproportionate downstream influence by reshaping daily routines, attentional priorities, and self-regulatory capacity.

In parallel, spillover and coaction theories posit that health behaviors are dynamically interconnected, such that initiating change in one domain can increase the probability of change in others ^16,17^. Rather than operating through direct causal transfer alone, these generalization effects are thought to emerge through multiple shared pathways ^13^. First, successful behavior change can mobilize common motivational and self-regulatory resources, including goal commitment, perceived control, and regulatory confidence, thereby lowering the psychological barriers to engaging in additional behaviors. Second, early success in one domain may enhance self-efficacy and outcome expectancies, reinforcing beliefs about one’s capacity to initiate and sustain health-related change more broadly. Third, behavior change can prompt shifts in health-related identity and self-concept, whereby individuals come to see themselves as “health-conscious” or “active agents,” increasing consistency pressures across behaviors. Finally, changes in daily routines, time allocation, and environmental contexts can restructure opportunity landscapes in ways that facilitate complementary behavior changes. Taken together, all these theoretical frameworks imply that health behaviors operate as interdependent components of a broader behavioral system, within which some behaviors may serve as leverage points capable of catalyzing wider lifestyle changes.

Empirical evidence remains mixed regarding which specific behaviors most reliably function as catalysts for broader lifestyle change. Physical activity has traditionally been positioned as the cornerstone of healthy lifestyles, supported by robust evidence linking regular activity to reduced cardiometabolic risk, improved mental health, and lower mortality, particularly among children, adolescents, and young adults ^18^. Consequently, many public health guidelines and intervention frameworks prioritize increasing physical activity as a primary prevention strategy. Nevertheless, evidence from intervention and spillover research suggests that gains in physical activity do not consistently generalize to improvements in other lifestyle domains, such as diet quality or sleep, and may be constrained by compensatory responses in energy intake or by the episodic and effortful nature of exercise engagement ^19,20^. In contrast, a growing body of research indicates that dietary behaviors may occupy a more foundational position in daily life. Eating behaviors are enacted multiple times per day, are deeply embedded in social routines and physical environments, and exert direct influence on physiological processes related to energy regulation, circadian rhythms, and affective functioning ^14,21,22^. Consistent with this view, diet quality has been linked to mental health outcomes, sleep quality, and cognitive functioning, and dietary interventions have demonstrated potential for downstream benefits beyond nutrition alone, including improvements in mood and overall wellbeing ^23,24^. From a self-regulatory and systems perspective, these characteristics suggest that dietary behaviors may function as keystone habits within broader lifestyle systems, with greater capacity to influence multiple behavioral pathways simultaneously than more episodic behaviors such as structured physical activity.

Capable of testing these possibilities through capturing conditional dependencies among multiple health behaviors simultaneously, the network approach of modeling treats behaviors as nodes connected by edges representing partial correlations among behaviors. When estimated with regularization and evaluated for accuracy and stability, these networks can reveal clusters, nodes linking clusters, and nodes with higher centrality that may be influential in the system ^25^. In mental health research, network theory has reframed psychopathology as a set of mutually reinforcing symptoms, helping to explain how specific symptoms (e.g., insomnia) propagate effects across the network ^26,27^, In behavior medicine and public health, similar logic can illuminate how lifestyle practices cohere and influence wellbeing.

We aimed to investigate the relative importance and interdependence pattern among diet, physical activity, sleep, social engagement, green/blue space exposure, substance use (alcohol and tobacco), using cross-sectional survey data collected from young Chinese adults. We adopted a network approach to testing our hypotheses that health behaviors form a robust network, wherein physical activity and/or diet are expected to be central nodes by exerting disproportionate influence over other health behaviors. All our hypotheses and analyses were pre-registration (https://aspredicted.org/5dxf-569m.pdf).

## Methods

### Design and Participants

We conducted a cross-sectional survey of health behaviors and emotional distress among Chinese college students from September to November 2024. The survey was split into two halves with the intention of allowing for a short break. Undergraduate students from multiple universities in China joined the research by using their cellphones to scan QR codes of the online survey, which were shared in messaging groups in WeChat application, on-campus handout distribution, and peer invitation. All students provided informed consent prior to responding to survey questions. The research procedures were reviewed and approved by the ethics committee of XXXXXXX (Approval #: H25145).

The currently reported data were collected from three universities in China and represented approximately a quarter of all the data collected due to access regulations imposed by Consortium of Physical and Mental health Promotion for Chinese Students (CPMC). To ensure data quality, we excluded cases that (a) included inappropriate responses on attention-check items, (b) associated with repeated responding attempts, (c) indicated highly unlikely demographic information (i.e., age < 16 or > 24; heights 3 SDs away from M), and (d) completed merely the first half of the survey. The final sample included 5,652 students and no missing responses. Figure 1 displays the geographic distribution of participants’ home residences across Chinese provinces and Table 1 summarizes demographic information of the sample.

**Figure 1.**
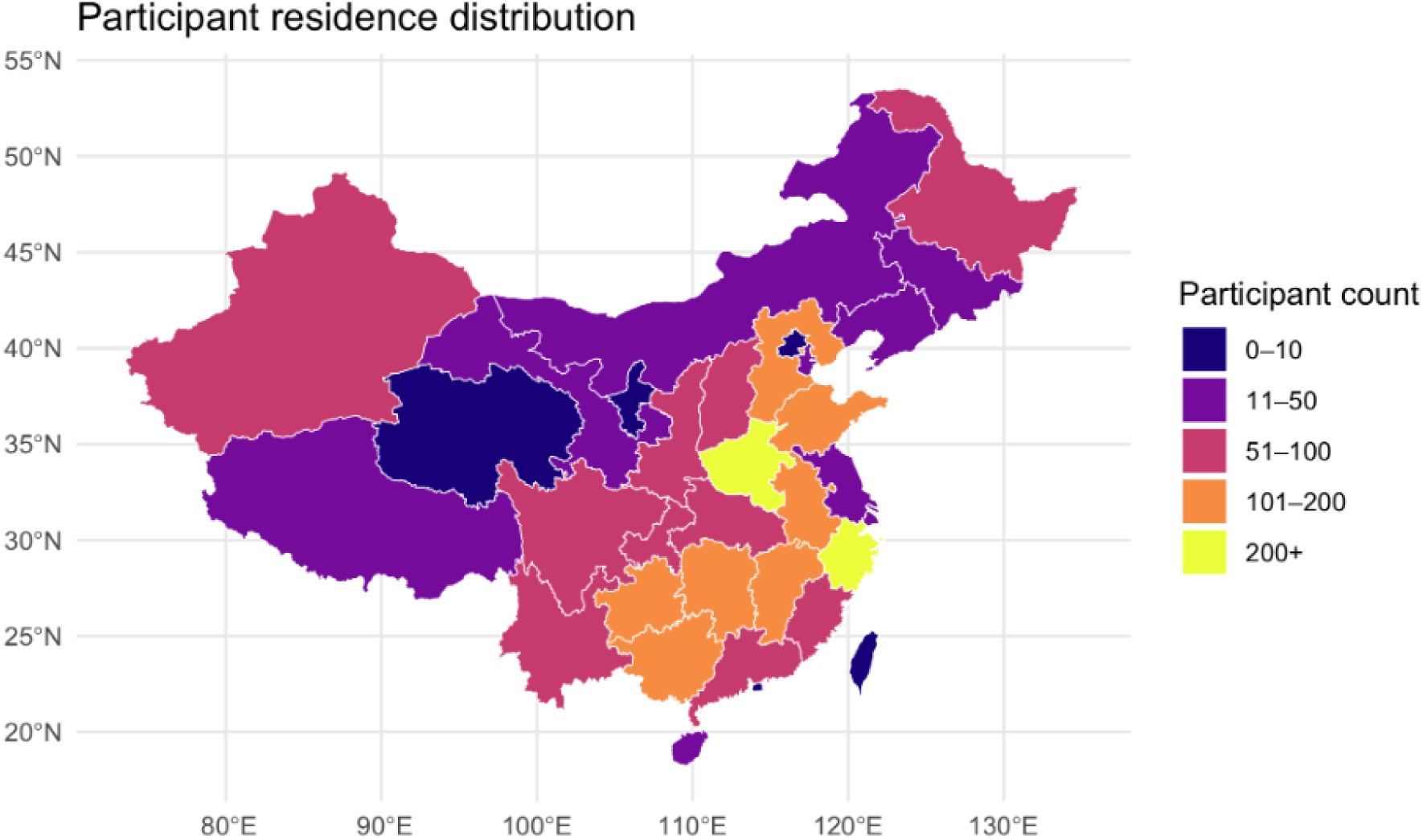
Distribution of participants’ home residences across provinces in China.

**Table 1.** Sample demographic information.

| Sample Characteristic | *n* (%) or *Mean* (*SD*) |
| --- | --- |
| Gender<sup>□</sup> |  |
| Male | 2375 (42.0) |
| Female | 3277 (58.0) |
| Class of Major in College<sup>□</sup> |  |
| Humanities | 2253 (39.9) |
| Natural Sciences | 3399 (60.1) |
| Being the Only Child<sup>□</sup> |  |
| Yes | 1704 (30.1) |
| No | 3948 (69.9) |
| Year in College<sup>□</sup> |  |
| First Year | 3191 (56.5) |
| Second Year | 2050 (36.3) |
| Third Year | 240 (4.2) |
| Fourth Year | 171 (3.0) |
| Residence Type Prior to Entering College<sup>□</sup> |  |
| Urban Area | 42.3 (2392) |
| Urban-Town Fringe | 1035 (18.3) |
| Town Area | 539 (9.5) |
| Town-Rural Fringe | 623 (11.0) |
| Rural Area | 1063 (18.8) |
| Paternal Education Level<sup>□</sup> |  |
| Primary School or Lower | 680 (12.0) |
| Middle School | 2100 (37.2) |
| High School | 1744 (30.9) |
| College or Higher | 1128 (20.0) |
| Maternal Education Level<sup>□</sup> |  |
| Primary School or Lower | 991 (17.5) |
| Middle School | 2028 (35.9) |
| High School | 1694 (30.0) |
| College or Higher | 939 (16.6) |
| Height (cm)<sup>□</sup> | 167.9 (8.3) |
| Weight (kg)<sup>□</sup> | 60.2 (12.3) |
| Age (year)<sup>□</sup> | 18.8 (1) |
| Body Mass Index (kg/m<sup>2</sup>)<sup>□</sup> | 21.2 (3.4) |

*Note.* <sup>a</sup> dichotomous variable. <sup>b</sup> ordinal variable. <sup>c</sup> continuous variable. Dichotomous and ordinal variables are summarized using percentage and count, *n* (%), and continuous variables using mean and standard deviation, *Mean* (*SD*). Whereas ordinal and continuous variables are modeled in the networks, dichotomous variables are used in network comparison tests.

### Measures

The survey focused on collecting one’s health behaviors and emotional distress. Seven health behaviors were measured using either established instruments or customary question items, including healthy diet, physical activity (International Physical Activity Questionnaire-Short Form [IPAQ-SF-7] and Physical Activity Rating Scale [PARS-3]), quality of sleep (Pittsburgh Sleep Quality Index [PSQI], Cronbach *α* = 0.762), social engagement (Social Network Questionnaire, Cronbach *α* = 0.832), drinking, and smoking. In addition, we measured symptoms of state anxiety (Generalized Anxiety Disorder [GAD-7], Cronbach *α* = 0.933) and depression (Patient Health Questionnaire [PHQ-9], Cronbach *α* = 0.896).

Rather than using established instrument, measurements of healthy diet, drinking, and smoking involved customary question items. We measured healthy diet using four items, including weekly frequency of having breakfast, weekly frequency of having dinner, intention to have healthy food for long-term health benefits, and intention to adopt Mediterranean-style diet. The dinner-frequency item was later dropped to enhance measurement reliability (Cronbach *α* = 0.76) and clarify interpretation (i.e., unlike having breakfast, having dinner may not indicate healthy diet among college students). For drinking, one question based on gender-specific criteria of Center of Disease Control (CDC) helped classify students into never-, non-, light, moderate, and heavy drinkers ^28^. Similarly, one question based on the Government of Canada helped classify students into never-, non-, light, moderate, and heavy smokers ^29^.

### Statistical Analysis

The preliminary analysis included computing the sum score (or total volume of physical activity from IPAQ-SF-7 and PARS-3) associated with each health behavior and emotional distress when measured with multiple question items. In addition, the interpretive direction of all the health behavior indicators was adjusted so that higher scores corresponded to healthier patterns of behavior. As both IPAQ-SF-7 and PARS-3 helped generate indicators of physical activity volume, we chose to focus on the indicator computed from IPAQ-SF-7 but performed sensitivity analysis for that from PARS-3, whose results remained consistent (see Appendix A). All continuous variables for health behaviors and emotional distress underwent nonparanormal transformation to enhance compliance of normality ^30^.

The pre-registered modeling approach of pairwise Markov random field helps understand the conditional dependencies in multivariate situations through partial correlations. With the EBICglasso estimator (*λ* = 0.50) that relies on graphical least absolute shrinkage and selection operator (glasso;^31^) and extended Bayesian information criterion (EBIC; *γ* = 0.50), resulting networks of clear structures by reducing spurious edges (i.e., partial correlations) among nodes (i.e., variables) in high-dimensional settings. When plotted with the Fruchterman-Reingold algorithm ^31^, nodes with more heavily weighted edges between each other are placed closer, and those with more edges to other nodes are placed closer to the center of the graph.

We specified three networks, including (1) health behavior network, (2) health behavior + emotional distress network, and (3) health behavior + emotional distress + demographic information network. Although all three network models help understand the interdependence among health behaviors, we focused on Network 3 given its inclusion of both emotional distress and demographic background and kept the other two networks as secondary models. To understand centrality of specific health behavior(s) in the network, we considered the indicator of betweenness (i.e., the total number of shortest paths passing the node), closeness (i.e., inverse of the distance between a node and all other nodes), and strength (i.e., sum of edge weights between a specific node and other connected nodes). Higher values on these indicators suggest higher centrality in the network.

We evaluated centrality stability using case-dropping bootstrapping with 5,000 resamples, reporting the proportion of sample subset to maintain a correlation-stability (CS) coefficient at the level of 0.70, which indicates the largest proportion of cases that can be removed while still maintaining, with 95% probability, a correlation of at least 0.70 between the original centrality indices and those obtained from networks estimated on the subsets. The CS coefficients should be 0.50 or higher to consider the centrality estimates stable ^25^. In addition, we assessed network edge-weight accuracy and tested edge weight differences with nonparametric bootstrapping involving 3,000 resamples ^25^.

Finally, we conducted Network Comparison Tests (NCT) with 1000 permutations to examine differences in network structure and global strength across gender, academic major, as well as freshman year vs. all other college years. For each comparison, we reported structure (M) and global strength (S) statistics with the permutation-based *p* values. Our interpretation of correlation coefficients is based on Cohen ^32^. All analyses were conducted in R with the *a* level set at 0.05.

## Results

Figure 2 displays the three specified network models, across which the connectivity among health behaviors remained visually consistent. Net 3 included 14 nodes and 51/91 non-zero edges (mean edge weight = 0.021), in comparison to Net 2 having 9 nodes and 33/36 non-zero edges (mean edge weight = 0.046) and Net 1 having 7 nodes and 20/21 non-zero edges (mean edge weight = 0.025). Visual inspections also suggested distinguishable clustering patterns, including a “positive lifestyle” cluster comprising diet, exercise, space, and social, a “mental distress and sleep quality” cluster that included sleep, anxiety, and depression, as well as a “substance use” cluster for drinking and smoking.

**Figure 2.**
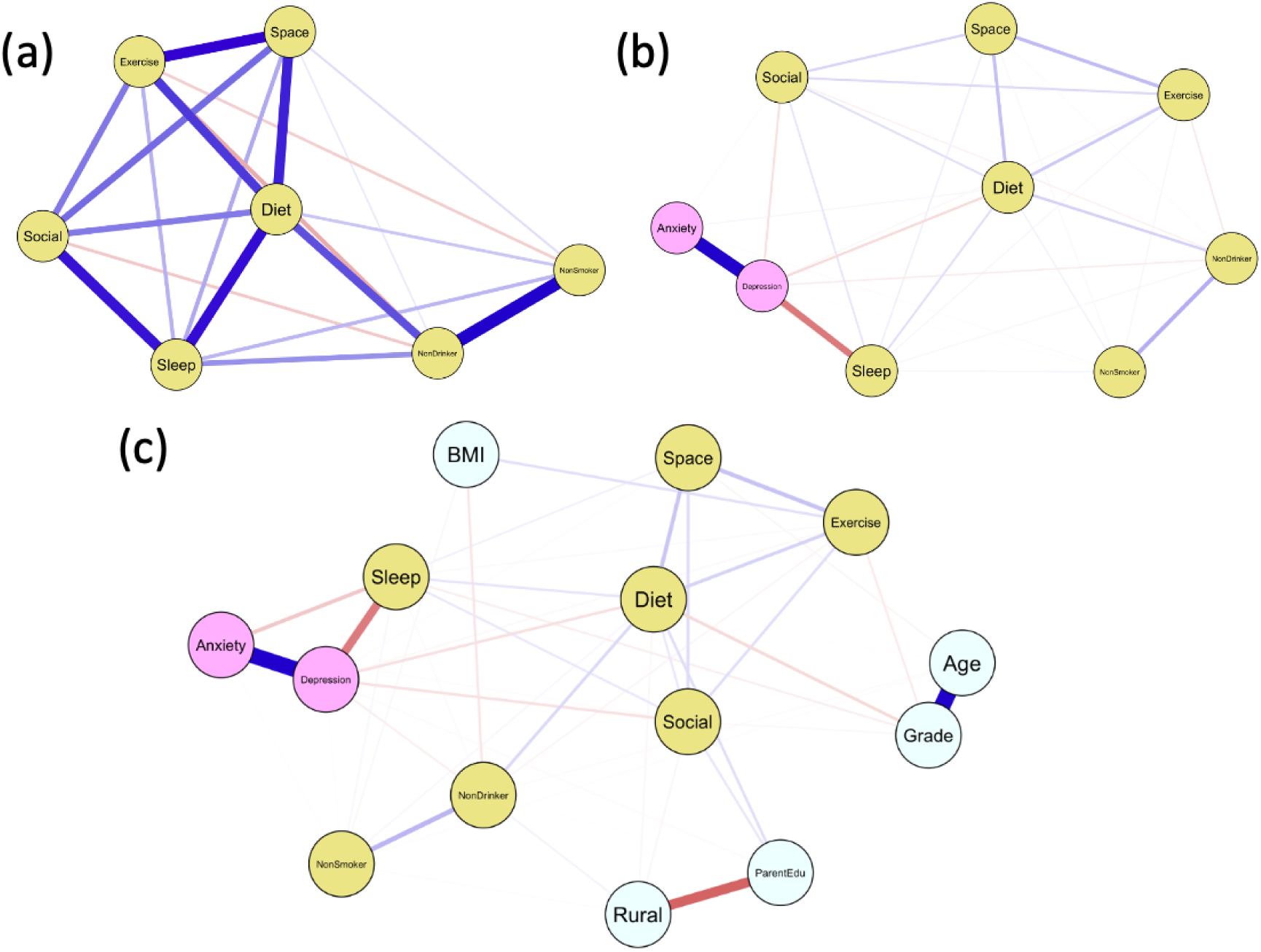
Between-subject network models comprising (a) health behavior only (Net 1), (b) health behavior + emotional wellbeing (Net 2), (c) health behavior + emotional wellbeing + demographic information (Net 3). Yellow nodes represent positive health behaviors, with pink and blue nodes correspond to emotional distress and demographic information, respectively. Exercise = transformed score of total volume of physical activity in metabolic equivalent of task (MET) based on IPAQ-SF-7; Diet = transformed sum score of weekly frequency of having a timely breakfast and intentions to engage in healthy diets; NonDrinker = habit of drinking alcohol with higher scores indicating lighter drinking; NonSmoker = habit of smoking with higher scores indicating lower volume of smoking; Sleep = the negative of transformed sum score of self-reported sleep problems based on PSQI; Social = transformed total number of relatives and online/offline friends for social engagement; Space = transformed monthly frequency of going to green (e.g., forest) and blue (e.g., lake, beach) space; Anxiety = transformed sum score of anxiety symptoms based on GAD-7; Depression = transformed sum score of depression symptoms based on PHQ-9; Age = student year of age; BMI = body mass index (kg/m^2^); Grade = year in college; ParentEdu = sum score of paternal and maternal education level; Rural = the residence location of core family with higher scores corresponding to more rural regions.

As Figure 3a illustrates, the node of diet, but not that of exercise, consistently demonstrated the centrality of betweenness (rb_net3_ = 53, rb_net2_ = 18, rb_net1_ = 8), closeness (rc_net3_ = 0.005, rc_net2_ = 0.011, rc_net1_ = 0.018), and strength (rs_net3_ = 0.694, rs_net2_ = 0.649, rs_net1_ = 0.665) among those of all health behaviors. Unlike diet whose strength centrality includes contributions from several edges, other nodes (e.g., anxiety, depression, sleep) demonstrating high strength centrality relied on specific edges that are heavily weighted. Figure 3b displays the CS coefficients for understanding stability of centrality indicators, showing that the centrality indicators in Figure 3a were stable estimates. In greater detail, the CS coefficients associated with betweenness (0.750, 0.594, 0.672), closeness (0.750, 0.750, 0.750), and strength (0.750, 0.750, 0.750 for Net 3, Net 2, and Net 1, respectively) were all larger than the recommended criterion of 0.50 with 0.750 as the maximum level of CS coefficient tested.

**Figure 3.**
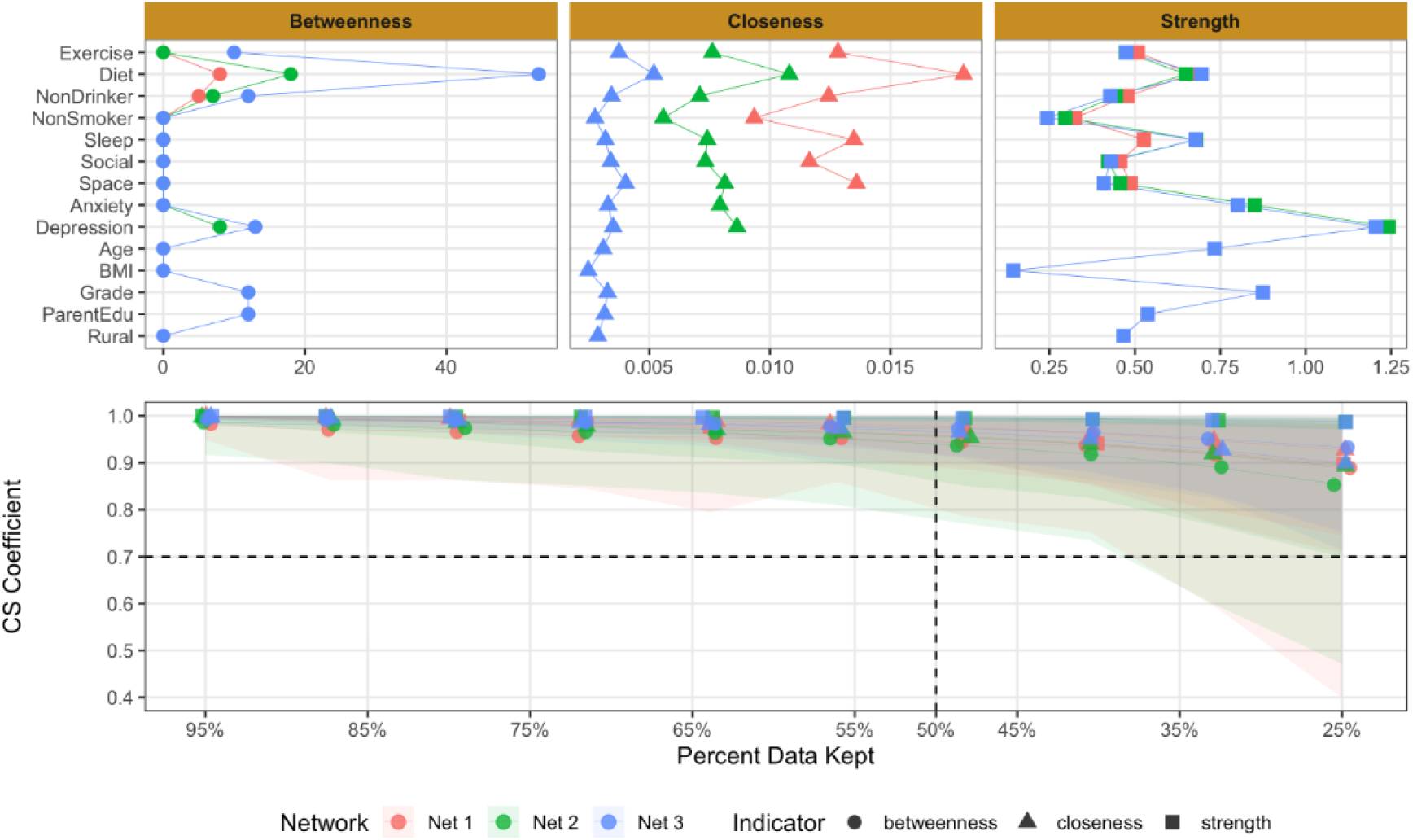
Network centrality indicators and their stability. Panel a displays four specific centrality indicators, which are represented using different point shapes, in all the three network models that are colored differently. Panel b illustrates the corresponding stability of each centrality indicator in Panel a based on case-dropping bootstraps. Note that, although the range of Panel a’s y-axis is [0, 1], we used [0.4, 1] instead with two reference lines (CS Correlation = 0.70, Percent Data Kept = 50%) to help interpret centrality stability based on Epskamp et al. (2018).

As Figure 4 shows, the centrality of the node of diet was also reflected by being meaningfully connected with other nodes of health behavior. In Net 1, diet positively correlated with exercise (0.125), space (0.143), and social (0.083), and negatively correlated with sleep (-0.153), drinking (-0.110), and smoking (-0.032). This pattern of correlation between diet and other health behaviors remained robust when emotional distress and demographic information was considered in modeling (i.e., Net 2 and Net 3). In the most inclusive Net 3, diet positively correlated with exercise (0.106), space (0.135), and social (0.054), and negatively correlated with sleep (-0.060), drinking (-0.094), and smoking (-0.024). Moreover, diet was positively correlated with parent education level (0.079), and negatively correlated with depression (-0.073) and grade (-0.089). Finally, edge difference tests indicated that diet-exercise had similar positive edge strength to diet-space (*p* > 0.05), both of which had more positive edge strength than diet-social (*p*s < 0.05), and diet-drinking had similar negative edge strength to diet-sleep (*p* > 0.05), both of which had more negative edge strength than diet-smoking (*p*s < 0.05).

**Figure 4.**
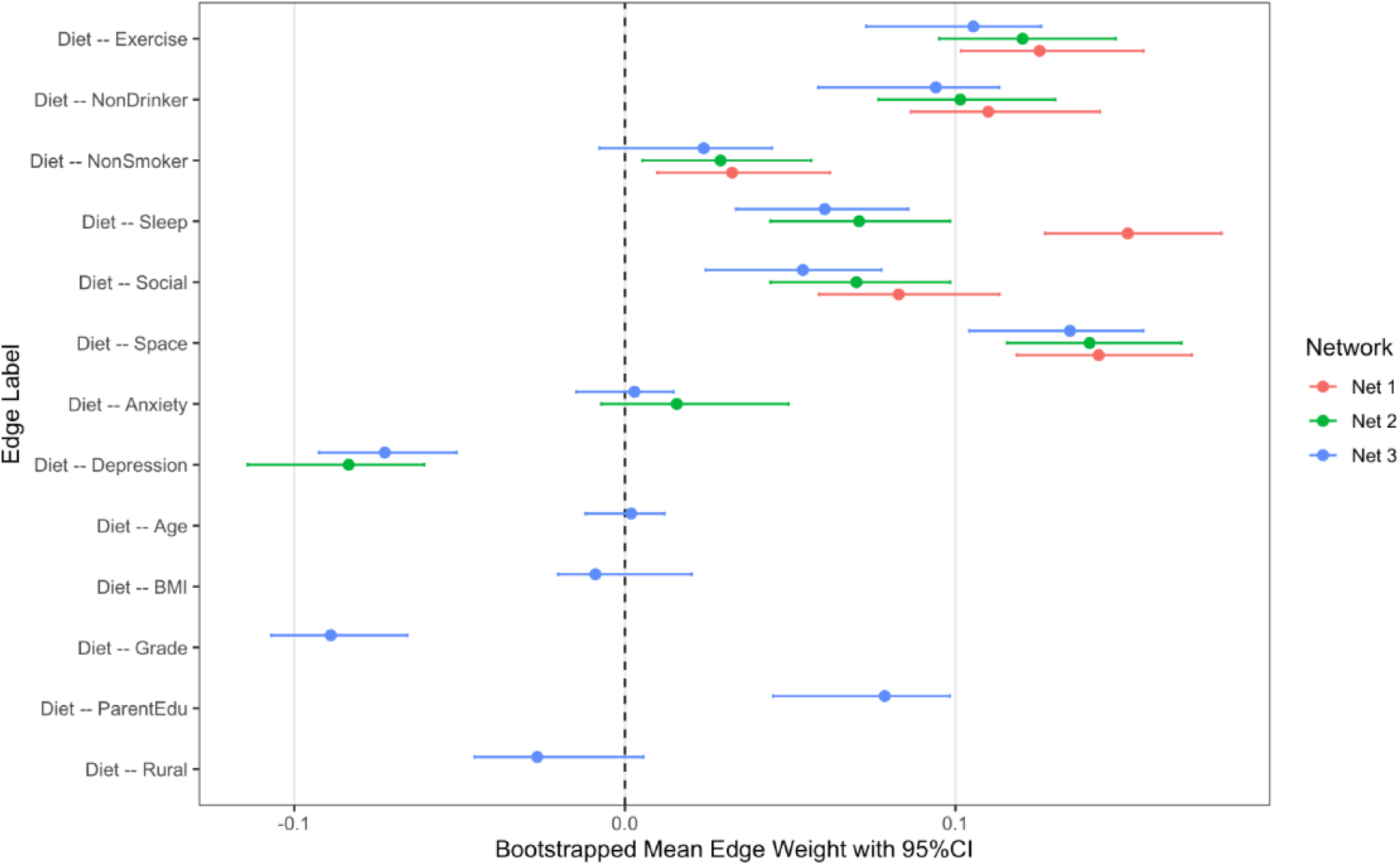
Bootstrapped mean edge weight with 95% bootstrap CI for diet-involved edges across all three network models.

Network comparison tests suggested no differences regarding network structure or overall strength for Net 3 when the sample was split and compared regarding gender (males vs. females: *M* = 0.062, *p* = 0.39, and *S* = 0.097, *p* = 0.73), academic major (humanities vs. natural sciences: *M* = 0.059, *p* = 0.53, *S* = 0.049, *p* = 0.87), and grade (freshman year vs. other college years: *M* = 0.041, *p* = 0.98, *S* = 0.112, *p* = 0.72), and consistent null results regarding network structure and overall strength were obtained for Net 1 and Net 2 (i.e., *p*s > 0.14).

## Discussion

In a large multi-university sample of Chinese college students, we mapped how everyday health behaviors interrelate with emotional distress and demographics using network analysis. Healthy diet, but not physical activity, emerged as the most central health behavior by exhibiting high and stable betweenness, closeness, and strength centrality in the behavior network. Such centrality of healthy diet was not the artifact of a single heavy edge. Instead, it reflected multiple moderate connections, positively to physical activity, green/blue-space exposure, and social engagement, as well as negatively to sleep problems and substance use (alcohol and tobacco). This pattern of networking persisted when emotional distress and demographic information was considered, and when key subgroups by gender, academic major, or year in college were compared. These findings point to diet as a structural hub within the lifestyle system of young Chinese adults.

The observed network structure reinforces the view that health behaviors operate as an integrated system rather than independent risk factors. Three clusters were consistently identifiable: a positive lifestyle cluster (diet, physical activity, social engagement, and green/blue-space exposure), a distress–sleep cluster (sleep problems, anxiety, and depression), and a substance use cluster (alcohol and tobacco). This organization aligns with ecological and biopsychosocial models, which posit that behaviors sharing similar contextual drivers and regulatory demands tend to co-activate ^12^. Notably, the positive lifestyle cluster illustrates how health-promoting behaviors may reinforce one another through shared routines and environments. Activities such as eating, socializing, and spending time in green or blue spaces often co-occur temporally and spatially, suggesting that interventions targeting one component may indirectly strengthen others. This provides empirical support for multi-behavior intervention strategies that emphasize lifestyle integration rather than isolated behavior change ^33^.

The centrality of diet, over and above physical activity, has important theoretical implications. From a self-regulation perspective, dietary behaviors are enacted frequently, require repeated decision-making, and are deeply embedded in daily schedules and social contexts. These characteristics make diet especially sensitive to environmental cues and capable of influencing downstream processes such as energy availability, sleep quality, and affect regulation. In contrast, physical activity, while critically important for health, is more episodic and may depend on upstream conditions such as nutrition, mood, and social support. From a behavioral contagion and spillover perspective, the findings suggest that dietary practices may serve as an efficient gateway for broader lifestyle change. Improvements in diet may enhance energy, mood, and self-efficacy, thereby increasing readiness for physical activity and social engagement, while simultaneously reducing reliance on maladaptive coping behaviors such as smoking or drinking. This asymmetry helps explain why interventions that successfully improve diet often yield secondary benefits in other domains ^34^, whereas exercise-only interventions sometimes show more limited generalization ^19^.

The clustering of sleep with anxiety and depression is consistent with network theories of mental health, which emphasize reciprocal reinforcement among symptoms and behaviors ^26^. Sleep appears to function as a bridge between emotional distress and daily lifestyle practices, reinforcing its value as an early indicator of broader psychosocial vulnerability in student populations. Monitoring sleep quality may therefore provide a practical screening tool for identifying individuals at risk for both mental health problems and deteriorating lifestyle patterns. Interestingly, alcohol and tobacco use formed a relatively peripheral cluster with weaker integration into the broader network. This pattern contrasts with findings from many Western samples, where substance use often co-occurs more strongly with poor diet, inactivity, and sleep disruption ^35^. The relative isolation of substance use in this sample may reflect cultural norm differences, lower prevalence rates among Chinese university students than western counterparts, or gendered social practices ^36–38^. This underscores the importance of cultural context when interpreting behavioral networks and cautions against assuming universality in clustering patterns according to the ecological perspective of health behaviors.

From an applied standpoint, the findings suggest that dietary behaviors represent a strategic intervention leverage point for improving overall student health. Campus-based initiatives that improve food environments, promote regular meals, or embed healthy eating within social and nature-based activities may yield cascading benefits across multiple behaviors. Such considerations are particularly relevant given the present observation that the degree of healthy diet tended to decline as college students became more senior (i.e., the correlation between diet and grade is -0.09). The observed invariance of network structure across gender, academic major, and year in college further suggests that diet-centered strategies may be broadly applicable and scalable within the Chinese higher education system. These implications align closely with national public health priorities, which emphasizes early prevention, lifestyle modification, and integrated health promotion. Rather than positioning diet and exercise as parallel but separate targets, the present findings argue for a systems-oriented design, with diet as an entry point supported by complementary changes in physical activity opportunities, social programming, and physical environments. For instance, a trial has shown the possibility of such system-oriented design by utilizing community gardening in a nature-based lifestyle intervention, wherein participants had healthy diets (e.g., diet high in fiber) while gardening together in green space, showed improvements in both dietary and physical-activity outcomes ^39^

### Limitations and Future Directions

Several limitations are worth noting for future endeavors. First, our cross-sectional, between-person network cannot establish temporal precedence or causal pathways because partial correlations reflect conditional associations instead of influence. Future work should use longitudinal designs and within-person network models to test directional influence, such as day-to-day spillovers, lagged effects (e.g., how diet affects sleep that night), and potential feedback loops.

Second, although most scales were validated, diet relied on custom items and one diet item was removed to improve reliability. Future work should create more standardized measurement tools with high external validity and triangulate it with self-reports to reduce potential bias from the measurement tool. For instance, recordings from mobile eye trackers during on-campus canteen purchase can be cross validated with dietary logs.

Finally, although our network comparison tests showed invariance by gender, major, and year in college among Chinese students, other moderators (socioeconomic status, residence, urbanicity) and contextual variables (food environment, green/blue-space access) may shape network structure and edge strength, particularly in samples from different non-Chinese cultures. Future work is encouraged to take a comparative perspective by incorporating cross-cultural or cross-regional samples, examining how environmental, socioeconomic, and policy factors influence the interrelations among health behaviors, and testing whether central nodes such as diet remain stable or shift across different cultural and ecological contexts.

## Conclusions

Taken together, this study demonstrates that health behaviors among young Chinese adults form a coherent and interpretable network in which healthy diet occupies a central, integrative position. By bridging physical activity, social engagement, environmental exposure, sleep, and substance use, diet emerges as a promising focal point for theory-driven, system-level interventions. These findings highlight the value of network approaches for advancing health behavior theory and for designing more effective, scalable health promotion strategies in university settings.

## Supporting information

eFigure1-3

## Data Availability

All data produced in the present work are contained in the manuscript

## Contributors

Concept and design: all authors

Acquisition, analysis, or interpretation of data: DW ZZ SL

Drafting of the manuscript: SL FH

Critical review of the manuscript for important intellectual content: GW DW ZZ

Statistical analysis: SL FH

Obtained funding: GW

Administrative, technical, or material support: LG FH

Supervision: GW SL

## Data Sharing Statement

No additional data are available.

## Acknowledgement

This work was supported by Consortium of Physical and Mental health Promotion for Chinese Students (CPMC).

## Supplementary Material (1)

Supplementary appendix

## References

1. Li X, Mohanty I, Zhai T, Chai P, Niyonsenga T. Catastrophic health expenditure and its association with socioeconomic status in China: evidence from the 2011-2018 China Health and Retirement Longitudinal Study. Int J Equity Health. 2023;22(1):194. doi:10.1186/s12939-023-02008-z

2. Fang Q, Ma G, Wang Y, et al. Current curative expenditure of non-communicable diseases changed in Dalian, China from 2017 to 2019: a study based on ‘System of Health Accounts 2011.’ BMJ Open. 2022;12(4):e056900. doi:10.1136/bmjopen-2021-056900

3. Xinhua. China has over 47 mln higher-education students in 2023. 2024. Accessed October 17, 2025. https://english.www.gov.cn/archive/statistics/202403/01/content_WS65e1d71dc6d0868f4e8e487a.html

4. Xinhua. China makes breakthrough in providing inclusive, high-quality education service: minister. 2025. Accessed November 17, 2025. https://english.www.gov.cn/news/202509/23/content_WS68d23513c6d00fa19f7a2a5d.html

5. Greene GW, Schembre SM, White AA, et al. Identifying Clusters of College Students at Elevated Health Risk Based on Eating and Exercise Behaviors and Psychosocial Determinants of Body Weight. J Am Diet Assoc. 2011;111(3):394–400. doi:10.1016/j.jada.2010.11.011

6. St-Onge MP, Grandner MA, Brown D, et al. Sleep Duration and Quality: Impact on Lifestyle Behaviors and Cardiometabolic Health. Circulation. 2016;134(18):e367–e386. doi:10.1161/CIR.0000000000000444

7. Doan N, Parker A, Rosati K, van Beers E, Ferro MA. Sleep duration and eating behaviours among adolescents: a scoping review. Health Promot Chronic Dis Prev Can Res Policy Pract. 2022;42(9):384–397. doi:10.24095/hpcdp.42.9.02

8. Twohig-Bennett C, Jones A. The health benefits of the great outdoors: A systematic review and meta-analysis of greenspace exposure and health outcomes. Environ Res. 2018;166:628–637. doi:10.1016/j.envres.2018.06.030

9. Gascon M, Zijlema W, Vert C, White MP, Nieuwenhuijsen MJ. Outdoor blue spaces, human health and well-being: A systematic review of quantitative studies. Int J Hyg Environ Health. 2017;220(8):1207–1221. doi:10.1016/j.ijheh.2017.08.004

10. WHO. Tackling NCDs: “best buys” and other recommended interventions for the prevention and control of noncommunicable diseases. 2017. Accessed October 17, 2025. https://www.who.int/publications/i/item/WHO-NMH-NVI-17.9

11. Mkhitaryan S, Crutzen R, Steenaart E, de Vries NK. Network approach in health behavior research: How can we explore new questions? Health Psychol Behav Med. 2019;7(1):362–384. doi:10.1080/21642850.2019.1682587

12. Sallis JF, Owen N. Ecological models of health behavior. In: Health Behavior: Theory, Research, and Practice, 5th *Ed*. Jossey-Bass/Wiley; 2015:43–64.

13. Wood W, Neal DT. A new look at habits and the habit-goal interface. Psychol Rev. 2007;114(4):843–863. doi:10.1037/0033-295X.114.4.843

14. van’t Riet J, Sijtsema SJ, Dagevos H, De Bruijn GJ. The importance of habits in eating behaviour. An overview and recommendations for future research. Appetite. 2011;57(3):585–596. doi:10.1016/j.appet.2011.07.010

15. Hennecke M, Bürgler S. Many roads lead to Rome: Self-regulatory strategies and their effects on self-control. Soc Personal Psychol Compass. 2020;14(6). doi:10.1111/spc3.12530

16. Dolan P, Galizzi MM. Like ripples on a pond: Behavioral spillovers and their implications for research and policy. J Econ Psychol. 2015;47:1–16. doi:10.1016/j.joep.2014.12.003

17. Johnson SS, Paiva AL, Mauriello L, Prochaska JO, Redding CA, Velicer WF. Coaction in Multiple Behavior Change Interventions: Consistency across Multiple Studies on Weight Management & Obesity Prevention. Health Psychol Off J Div Health Psychol Am Psychol Assoc. 2014;33(5):475–480. doi:10.1037/a0034215

18. Bull FC, Al-Ansari SS, Biddle S, et al. World Health Organization 2020 guidelines on physical activity and sedentary behaviour. Br J Sports Med. 2020;54(24):1451–1462. doi:10.1136/bjsports-2020-102955

19. Feng J, Huang WY, Zheng C, et al. The Overflow Effects of Movement Behaviour Change Interventions for Children and Adolescents: A Systematic Review and Meta-Analysis of Randomised Controlled Trials. Sports Med Auckl Nz. 2024;54(12):3151–3167. doi:10.1007/s40279-024-02113-1

20. Beaulieu K, Blundell JE, van Baak MA, et al. Effect of exercise training interventions on energy intake and appetite control in adults with overweight or obesity: A systematic review and meta-analysis. Obes Rev. 2021;22(S4):e13251. doi:10.1111/obr.13251

21. Story M, Kaphingst KM, Robinson-O’Brien R, Glanz K. Creating Healthy Food and Eating Environments: Policy and Environmental Approaches. Annu Rev Public Health. 2008;29(1):253–272. doi:10.1146/annurev.publhealth.29.020907.090926

22. Lassale C, Batty GD, Baghdadli A, et al. Healthy dietary indices and risk of depressive outcomes: a systematic review and meta-analysis of observational studies. Mol Psychiatry. 2019;24(7):965–986. doi:10.1038/s41380-018-0237-8

23. Godos J, Grosso G, Castellano S, Galvano F, Caraci F, Ferri R. Association between diet and sleep quality: A systematic review. Sleep Med Rev. 2021;57:101430. doi:10.1016/j.smrv.2021.101430

24. Solomou S, Logue J, Reilly S, Perez-Algorta G. A systematic review of the association of diet quality with the mental health of university students: implications in health education practice. Health Educ Res. 2023;38(1):28–68. doi:10.1093/her/cyac035

25. Epskamp S, Borsboom D, Fried EI. Estimating psychological networks and their accuracy: A tutorial paper. Behav Res Methods. 2018;50(1):195–212. doi:10.3758/s13428-017-0862-1

26. Borsboom D. A network theory of mental disorders. World Psychiatry. 2017;16(1):5–13. doi:10.1002/wps.20375

27. Robinaugh DJ, Hoekstra RHA, Toner ER, Borsboom D. The Network Approach to Psychopathology: A Review of the Literature 2008–2018 and an Agenda for Future Research. Psychol Med. 2020;50(3):353–366. doi:10.1017/S0033291719003404

28. CDC. NHIS - Adult Alcohol Use - Glossary. November 14, 2024. Accessed October 17, 2025. https://archive.cdc.gov/www_cdc_gov/nchs/nhis/alcohol/alcohol_glossary.htm

29. Canada H. Canadian tobacco use monitoring survey: Terminology. May 1, 2005. Accessed October 17, 2025. https://www.canada.ca/en/health-canada/services/health-concerns/tobacco/research/tobacco-use-statistics/terminology.html

30. Liu H, Han F, Yuan M, Lafferty J, Wasserman L. High-dimensional semiparametric Gaussian copula graphical models. Ann Stat. 2012;40(4):2293–2326. doi:10.1214/12-AOS1037

31. Friedman J, Hastie T, Tibshirani R. Sparse inverse covariance estimation with the graphical lasso. Biostatistics. 2008;9(3):432–441. doi:10.1093/biostatistics/kxm045

32. Cohen J. Statistical Power Analysis for the Behavioral Sciences. 2nd ed. Routledge; 2013. doi:10.4324/9780203771587

33. Zhang AL, Liu S, White BX, et al. Health-promotion interventions targeting multiple behaviors: A meta-analytic review of general and behavior-specific processes of change. Psychol Bull. 2024;150(7):798–838. doi:10.1037/bul0000427

34. Bayes J, Schloss J, Sibbritt D. The effect of a Mediterranean diet on the symptoms of depression in young males (the “AMMEND: A Mediterranean Diet in MEN with Depression” study): a randomized controlled trial. Am J Clin Nutr. 2022;116(2):572–580. doi:10.1093/ajcn/nqac106

35. de Vries H, van ‘t Riet J, Spigt M, et al. Clusters of lifestyle behaviors: Results from the Dutch SMILE study. Prev Med. 2008;46(3):203–208. doi:10.1016/j.ypmed.2007.08.005

36. Xie H, Di X, Liu S, Zeng X, Meng Z, Xiao L. Tobacco Use and Cessation Among College Students — China, 2021. China CDC Wkly. 2022;4(21):448–451. doi:10.46234/ccdcw2022.100

37. NIAAA. College Drinking Prevention. 2024. Accessed October 17, 2025. https://www.collegedrinkingprevention.gov/

38. ACHA. American College Health Association 2023. 2023. Accessed October 17, 2025. https://www.health-mental.org/american-college-health-association-2023/

39. Litt JS, Alaimo K, Harrall KK, et al. Effects of a community gardening intervention on diet, physical activity, and anthropometry outcomes in the USA (CAPS): an observer-blind, randomised controlled trial. Lancet Planet Health. 2023;7(1):e23–e32. doi:10.1016/S2542-5196(22)00303-5

