## Supplementary material for "Healthy Diet is Central in the Network of Health Behaviors among Young Chinese Adults": eFigure1-3

**Supplementary Online Content**

eFigure1. Between-subject network models comprising (a) health behavior only (Net 1), (b) health behavior + emotional wellbeing (Net 2), (c) health behavior + emotional wellbeing + demographic information (Net 3).

eFigure2. Network centrality indicators and their stability.

eFigure3. Bootstrapped mean edge weights with 95% bootstrap CI’s for diet-involved edges across all three network models.

eFigure1. Between-subject network models comprising (a) health behavior only (Net 1), (b) health behavior + emotional wellbeing (Net 2), (c) health behavior + emotional wellbeing + demographic information (Net 3). Yellow nodes represent positive health behaviors, with pink and blue nodes correspond to emotional distress and demographic information, respectively. Exercise = transformed score of total volume of physical activity in metabolic equivalent of task (MET) based on PARS-3; Diet = transformed sum score of weekly frequency of having a timely breakfast and intentions to engage in healthy diets; NonDrinker = habit of drinking alcohol with higher scores indicating lighter drinking; NonSmoker = habit of smoking with higher scores indicating lower volume of smoking; Sleep = the negative of transformed sum score of self-reported sleep problems based on PSQI; Social = transformed total number of relatives and online/offline friends for social engagement; Space = transformed monthly frequency of going to green (e.g., forest) and blue (e.g., lake, beach) space; Anxiety = transformed sum score of anxiety symptoms based on GAD-7; Depression = transformed sum score of depression symptoms based on PHQ-9; Age = student year of age; BMI = body mass index (kg/m^2^); Grade = year in college; ParentEdu = sum score of paternal and maternal education level; Rural = the residence location of core family with higher scores corresponding to more rural regions.

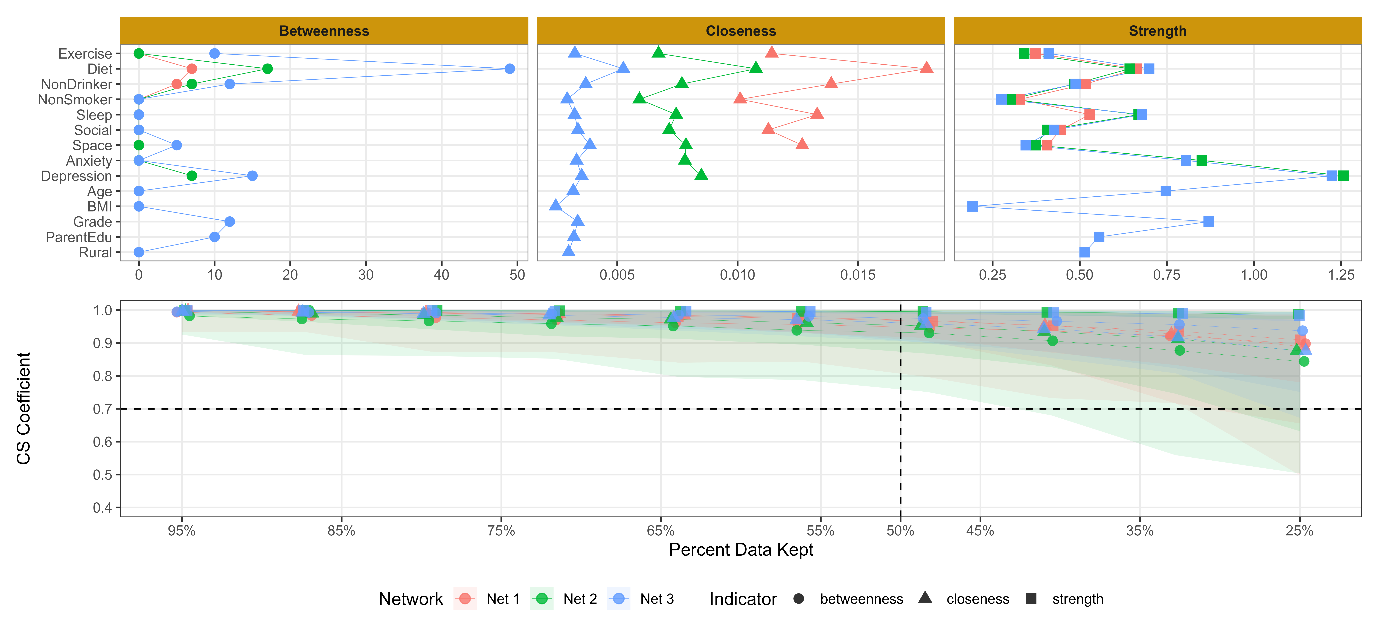

eFigure2. Network centrality indicators and their stability. Panel a displays four specific centrality indicators, which are represented using different point shapes, in all the three network models that are colored differently. Panel b illustrates the corresponding stability of each centrality indicator in Panel a based on case-dropping bootstraps. Note that, although the range of Panel a’s y-axis is [0, 1], we used [0.4, 1] instead with two reference lines (CS Correlation = 0.70 , Percent Data Kept = 50%) to help interpret centrality stability based on Epskamp et al. (2018).

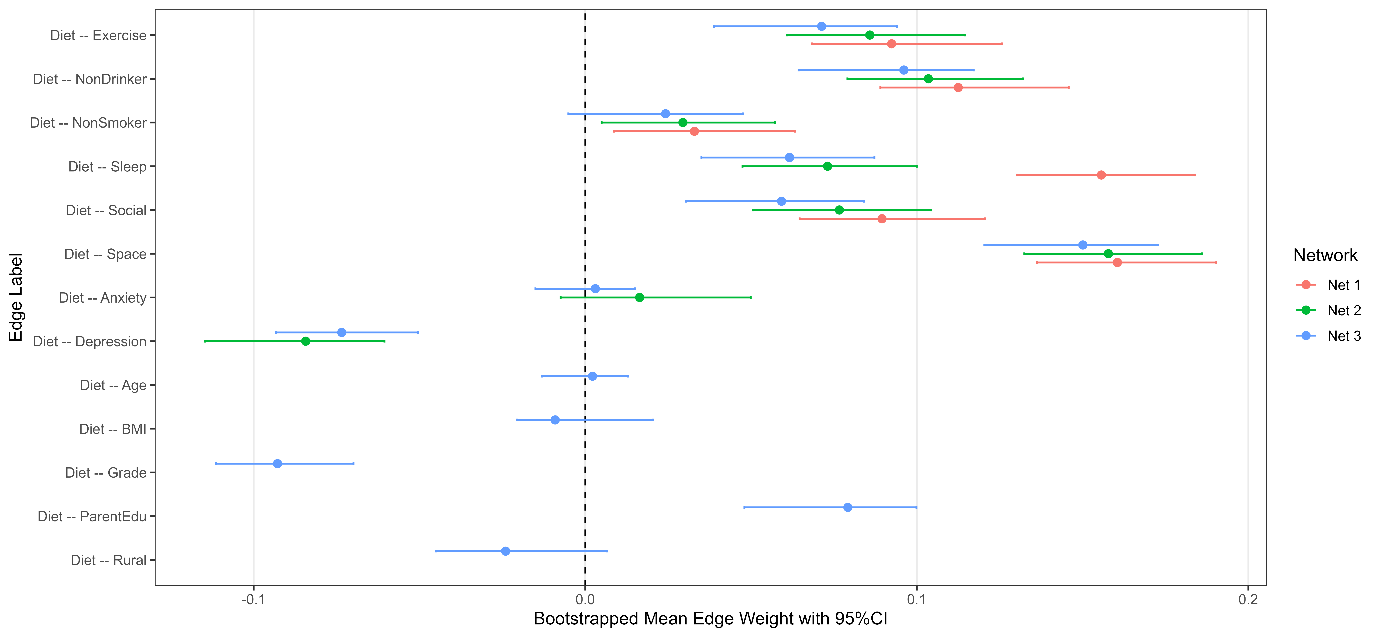

eFigure3. Bootstrapped mean edge weights with 95% bootstrap CI’s for diet-involved edges across all three network models.
